# Americans in rural areas are less likely to receive stress management counseling from physicians: a national repeated cross-sectional study of the 2018 and 2019 National Ambulatory Medical Care Survey

**DOI:** 10.1101/2024.09.24.24313205

**Authors:** Stephanie Marie Ira, Chris Gillette

**Author notes:** Address correspondence to: Chris Gillette, PhD, Wake Forest School of Medicine, Department of PA Studies, Medical Center Blvd., Winston-Salem, NC 27157.

## Abstract

Stress is a significant contributor to suicide ideation and attempts. This paper (1) describes the frequency of clinic visits in which physicians record stress management counseling; and (2) identifies visit-, physician-, and patient-related predictors of stress management counseling. We conducted a secondary analysis of the 2018 and 2019 National Ambulatory Medical Care Survey (NAMCS) clinic visit datasets. We identified clinic-, patient-, and physician-related predictors of stress management counseling. All analyses used weighted adjustment to account for the complex survey design. The weighted sample included 1,495,326,615 visits (unweighted (n=14,175) in 2018-2019. Combined, 1.32 per 100 visits (95% CI=0.72-1.92) included stress management counseling. Stress management counseling was less likely to occur when the visit occurred in a rural area (OR=0.22, 95% CI=0.09-0.53), among other characteristics. Stress management counseling rarely occurs during physician office visits in rural areas, despite rural areas suffering disproportionately worse health outcomes than urban areas.

## INTRODUCTION

Although stress can be a normal, healthy part of life, 77% of Americans say stress affects their physical health according to the American Psychological Association [1]. This chronic exposure to stress hormones causes the body to be more susceptible to adverse health events, and can affect the cardiovascular, digestive and immune systems [2-3]. Stress is a well-known factor that increases risk for suicide ideation and attempts [4]. Further, it is becoming well-established that social drivers of health and adverse childhood experiences are key drivers of racial/ethnic health disparities as well as physical and mental health, mainly because of longitudinal and disproportionate exposure to stress [5-6].

Additionally, Americans in rural areas experience greater morbidity and mortality compared to their urban counterparts, especially regarding mental health. Prevalence of mental health conditions is similar between rural and urban areas in the United States; however, those who live in rural areas are less likely than those in urban areas to have access to a mental health care provider. Americans who live in rural areas are also more likely to die by suicide than those in urban areas. Despite this well-known morbidity/mortality mental health disparities in rural areas, little is known about how physicians in ambulatory settings discuss mental health and stress.

A previous study of physician provision of stress management counseling was published using data from the National Ambulatory Medical Care Survey (NAMCS) between 2006 through 2009 [6]. This study found that only 3% of physician visits included stress management counseling, which was far lower than other health education/counseling areas (e.g., nutrition, diet/exercise). While the most recent national examination of stress management counseling by physicians used data from almost 15-20 years ago, many studies have identified the increasing importance of stress on negatively impacting health outcomes. Given the increasing recognition of the role of stress on health and racial/ethnic health disparities, it is important to provide an update of stress management counseling during physician visits. It is also important to understand the current practices in who is receiving stress management counseling and what factors are associated with its provision in order to design future interventions to improve the rates at which physicians discuss stress with their patients. Therefore, the objectives of this study are to: (1) describe the percentage of clinic visits in which physicians record stress management counseling; (2) identify visit- and patient-related predictors that are associated with provision of stress management counseling.

## METHODS

### Overview and Study Design

The Wake Forest University School of Medicine Institutional Review Board approved this study. This study was a secondary analysis of the 2018 and 2019 National Ambulatory Medical Care Survey (NAMCS). We combined two years to analyze because the 2019 NAMCS survey results regarding stress management counseling had a relative standard error (RSE) greater than 30%, indicating unreliable estimates regarding the primary outcome, even though there were more than 30 observed instances. Incorporating the 2018 dataset reduced the RSE to less than 30% (23.42%), indicating that the estimates were reliable, consistent with guidance from the National Center for Health Statistics’ NAMCS use. We excluded pre- and post-operative clinic visits as well as visits for children and adolescents younger than 17 years of age.

The NAMCS datasets have been well described [11]. Briefly, NAMCS is an annual survey that is conducted by the National Center for Health Statistics (NCHS) for non-federal physician visits. Prior to 2016, more than 99% of the data were collected by Census Field Representatives in conjunction with physicians and staff at sampled clinics. In 2018 and 2019, all data were collected by Census Field Representatives. In-depth explanations and coding for each NAMCS variable can be found online [12].

### Measures

*A priori*, we used Andersen’s Behavioral Model of Health Services Utilization (ABM) to guide selection of variables [13]. The ABM posits that access to health care is a function of predisposing characteristics (e.g., age, gender identity), enabling characteristics (e.g., having health insurance), and need characteristics (e.g., number of health conditions). We operationalized each construct of the ABM and then examined NAMCS documentation to identify and select only those variables for analysis. **Figure 1** contains the operationalization and the selected variables. For descriptive purposes, we used the separate race and ethnicity variables to describe patient visits but to ensure accuracy regarding race and ethnicity variables in the analyses, we only used the imputed race/ethnicity variable.

### Measures, Predisposing Factors

We measured patient age using categories (15-24 years, 25-44, 45-64, 65-74, 75 years and over). Patient sex is measured dichotomously (1=female, 2=male). Patient racial/ethnic identity is measured categorically (1=Non-Hispanic White, 2=Non-Hispanic Black, 3=Hispanic, 4=Non-Hispanic Other).

### Measures, Enabling Factors

The authors selected the following categorical variables from the dataset: Source of payment (All sources blank, unknown, private insurance, Medicare, Medicaid, CHIP, other state program, Worker’s compensation, self-pay, no charge/charity, other), whether the physician seeing the patient was the patient’s primary care physician is measured categorically (1=yes, 2=no, - 8=unknown, -9=blank), physician specialty (1=primary care, 2=surgical care, 3=medical care), whether the clinic was a solo or group practice (1=solo, 2=non-solo, -6=refused to answer, - 8=unknown, -9=blank), clinic ownership (1=physician/physician group, 2=medical/academic health center; community health center; other hospital, 3=insurance company, health plan, or HMO; other health corporation; other, -6=refused to answer, -8=unknown, -9=blank),. The following dichotomous variables (yes v. no) were selected: (1) Whether the patient was an established patient is measured dichotomously, whether a physician assistant (PA) was involved in the visit, whether a nurse practitioner (NP) was involved in the visit, whether another type of clinician involved in the visit, whether the physician was an allopathic (MD) physician or an osteopathic physician (DO), and whether the clinic was located in an rural area (1=yes, 0=no).

### Measures, Need Factors

The authors selected the following categorical variables from the dataset for the need factors: major reason for visit (1=new problem (<3 months onset), 2=chronic problem, routine, 3=chronic problem, flare-up, 6=preventive care, -9=blank), and whether the visit was related to injury/trauma or adverse effect of medical/surgical treatment (1=Yes, injury/trauma, 2=yes, overdose/poisoning, 3=yes, adverse effect of medical/surgical treatment, 4=no, 5=questionable injury status, -8=unknown, -9=blank). The following continuous variables were selected and analyzed for this study: number of medications discussed, total number of chronic conditions, number of areas counseled, total number of services ordered/provided, including vital signs, and time spent with the physician during the visit (minutes). The authors also identified whether the patient currently has cancer (yes v. no). The following dichotomous (yes v. no) health education/counseling variables were also selected: whether an asthma action plan was given, asthma education, growth development, sexually transmitted infection prevention, genetic counseling, alcohol abuse, substance abuse, family planning/contraception, stress management, injury prevention, diabetes education, tobacco use/exposure, weight reduction, exercise, and diet/nutrition.

### Statistical Analysis

All analyses were conducted using SAS v9.4 (Cary, NC). First, we describe the prevalence of visits that included stress management counseling per 100 visits. Second, we describe the characteristics of visits that included stress management counseling compared to visits that did not include stress management counseling using appropriate weighted bivariate statistical tests. We then constructed weighted multivariable logit models using the statistically significant variables from the bivariate tests to examine predictors for stress management counseling. The weighted logit models were two-sided with an α < 0.05 to indicate statistical significance and adjusted for patient complexity. Patient complexity was defined as the number of chronic conditions and the number of services that were provided/ordered. Results from the weighted logit models are presented using odds ratios (OR) and 95% Confidence Intervals (CI).

## RESULTS

There were 14,175 included observed visits, representing 1,495,326,615 non-federal physician visits during 2018 and 2019. In 2018, stress management counseling occurred in 1.22 visits per 100 visits (95% CI=0.63-1.81) while in 2019, stress management counseling occurred in 1.39 visits per 100 (95% CI=0.42-2.36) (Rao-Scott χ^2^ = 0.09, p=0.76). Over the two combined years, 1.32 visits per 100 visits (95% CI=0.72-1.92) included stress management counseling. **Table 1** presents the visit characteristics for visits in which stress management counseling is provided compared to visits in which stress management counseling was not provided.

The patient education and counseling descriptive statistics are presented in **Table 2** for the entire sample as well as primary care visits. Stress management was only discussed 1.3% of all visits and 2.09% of primary care visits. Of visits that included stress management discussions (n=220), the most commonly discussed areas were diet/exercise, nutrition, and weight reduction. The mode, median, 25^th^ and 75^th^ percentiles of the number of areas discussed in visits were all 0. The majority of visits (85.35%, n=12,099 unweighted, n=1,215,807,678) did not have any health education/counseling. There were no statistically significant patient-specific factors (race, sex, payment type) that were associated with whether a visit had any health education and counseling (n=2,076 unweighted, n=279,518,938, results not shown, all p>0.05).

**Table 3** presents the weighted logit regression model that was adjusted for patient complexity. This model found that stress management counseling was less likely to occur when: (1) when the visit occurred in a rural area (OR=0.22, 95% CI=0.09-0.53), (2) when an ‘other’ type of clinician was involved in the visit (OR=0.30, 95% CI=0.12-0.77), (3) when the physician was a surgeon (OR=0.06, 95% CI=0.01-0.29), and (4) when a physician or physician group owned the clinic compared to an academic medical center/community health center/other hospital (OR=0.23, 95% CI=0.09-0.59). The model also found that stress management counseling was more likely to occur when: (1) the visit occurred in solo physician offices (OR=3.29, 95% CI=1.39-7.81), (2) when the physician counseled on more areas (OR=2.10, 95% CI=1.60-2.75), and (3) when the clinic’s ownership type was not recorded compared to medical/academic health center, community health center, other hospital type (OR=4.03, (1.29-12.61).

## DISCUSSION

To our knowledge, this study contains the most recent examination of stress management counseling by physicians using nationally representative data. We found that approximately 1% of ambulatory office visits included discussions about stress management. We also found that physicians who practice in rural areas are less likely to provide stress management counseling than physicians in urban areas. This is an important practice gap that potentially might impact mental health outcomes for rural Americans. For those who wish to increase stress management counseling, we found that physicians were more likely to educate/counsel patients about stress when they educate about other areas, such as diet and exercise, suggesting that physicians combine multiple areas of patient education during clinic visits.

Compared to the most recent examination of stress management counseling that we are aware of (2006-2009 NAMCS data), we found that stress management was no longer the least common patient education area discussed, which is a positive finding [6]. Compared to that previous study from 2006-2009, we found the percent of visits in which physicians discussed stress management decreased (3% of visits compared to <2% of visits). Nerurkar and colleagues stated, in 2013, that physician counseling about stress had not been incorporated into primary care visits as much as other types of health education, such as diet/nutrition and exercise. This makes sense because the United States Preventive Services Task Force (USPSTF) guidelines recommend that physicians screen for diabetes and discuss weight loss to prevent obesity, both of which carry a Grade ‘B’ recommendation [14]. Stress management is not specifically mentioned by the USPSTF, which may be the reason why stress management has not been integrated by physicians as much as other counseling areas. The recent USPSTF recommendation to screen all patients for anxiety may offer an opportunity for physicians to integrate stress management into clinic visits more effectively [15]. Given the large literature on how stressful life events impacts depression, obesity, smoking, cardiovascular disease, racial/ethnic health disparities, and suicide, it is important for physicians to discuss stress with all patients [15-17]. Future research needs to investigate why stress management rates seem to have declined over time.

The major finding from this study is that stress management was about 79% less likely to occur when the clinic visit occurred in a rural area. It is well-known that Americans who live in rural areas experience mental health care disparities mostly stemming from lack of access to mental health specialists, and that these disparities are worsening over time [11, 18-19]. Research consistently shows that rural areas experience higher suicide rates than urban areas – double for teens as compared to urban areas and as four times higher in adults [19-20]. Stress is a key risk factor for suicidal ideation and mental health morbidity/mortality. Since rural areas tend to have less access to mental health care along with greater stigmatization of mental health, the role of the PCP is crucial to reducing the stigma of mental health in rural areas as well as providing more direct mental health care. Primary care has long been called the ‘de facto mental health care system’ and it is past time for medical educators and primary care training organizations to update PCPs’ training to provide more mental health care. The fact that visits in rural areas tended to have fewer visits in which physicians counseled about stress management is a key disparity that needs to be addressed.

One small previous study demonstrated that short, cognitive–behavioral stress management training reduces stress response as measured in saliva to an acute stressor in healthy male subjects – suggesting that effective stress counseling can be impactful in a clinic visit alone [22]. This data along with our findings supports clinicians in adding stress counseling into their visits to effect change in their patients’ mental and physical health.

There were no statistically significant patient-specific factors (race, sex, payment type) that were associated with whether a visit had any health education and counseling. This is encouraging from a patient-centered view of care, suggesting that all patients have an equal opportunity to receive stress management counseling. However, given the pervasive sociohistorical structural racism that Black, Hispanic/Latino, and American Indian/Alaska Native have endured for centuries in the United States and the current mental health disparities, it is important for physicians to address stress management with individuals in these populations.

Like other studies, this study has limitations. First, NAMCS only samples office-based physicians who are not employed by the federal government, so our results may not be indicative of all types of ambulatory clinic visits in the United States. Second, the NAMCS only samples physician visits which means that our findings may not reflect PA/NP counseling about stress management. Third, the NAMCS only samples one patient visit in time and visits in which stress may be discussed may not have been captured during the sampled visits (e.g., a regular well visit).

In conclusion, only 1.3% of outpatient clinic visits for adults in 2018 and 2019 included counseling on stress, yet studies routinely show that stress negatively affects mental and physical health. There is an extraordinary need to implement interventions to increase stress management counseling during routine clinic visits to mitigate the downstream physiologic and behavioral effects of stress.

## Supporting information

STROBE checklist

## Data Availability

All data produced are available upon reasonable request to the authors.

## Acknowledgements

This study did not receive any funding. The authors report no conflicts of interest.

