## Supplementary figures and images for "Americans in rural areas are less likely to receive stress management counseling from physicians: a national repeated cross-sectional study of the 2018 and 2019 National Ambulatory Medical Care Survey"

### Figure

**Figure 1: Andersen Behavioral Model Operationalization**

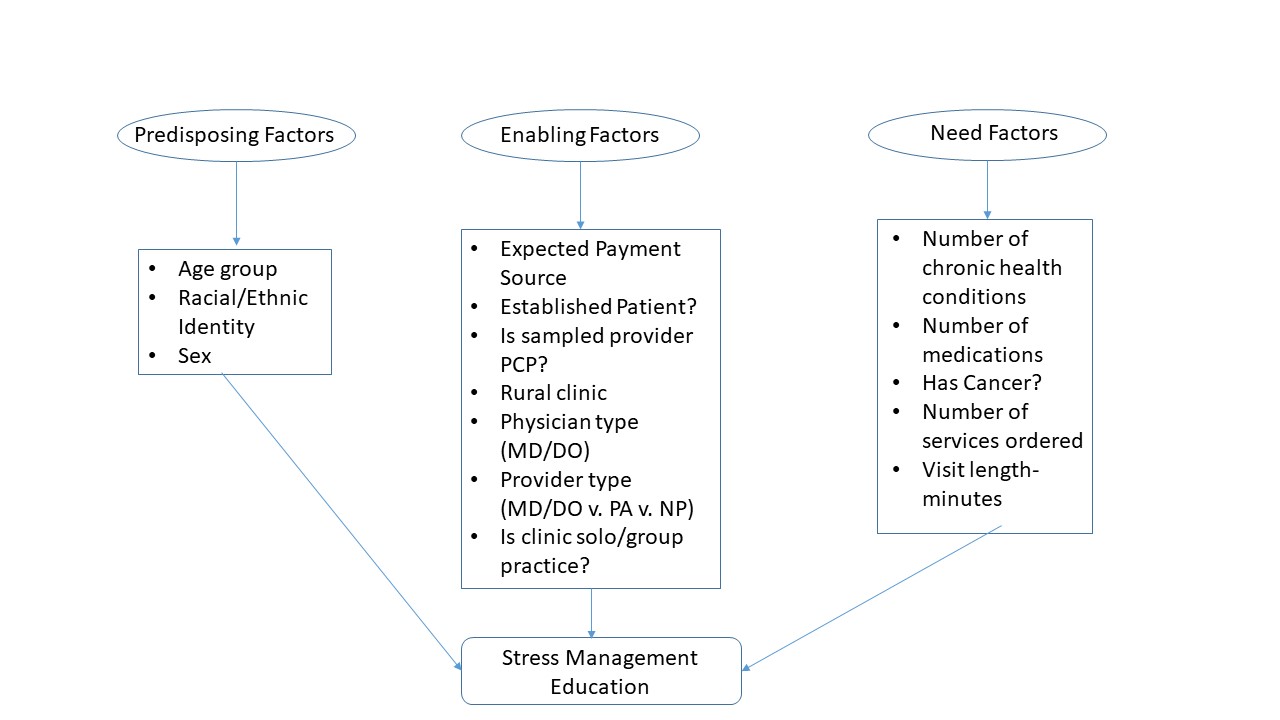
