## Supplementary material for "Americans in rural areas are less likely to receive stress management counseling from physicians: a national repeated cross-sectional study of the 2018 and 2019 National Ambulatory Medical Care Survey": Tables

**Table 1: Descriptive Statistics of Stress Management Discussions in Clinic visits (weighted n=1,495,326,615, unweighted=14,175)**

| **Characteristic** | **Stress Not Discussed During Clinic Visit (wgt n=1,475,637,700, unwght n=13,955) Percent (n)** | **Weighted Percent** | **Unweighted n** | **Stress Discussed During Clinic Visit (wght n=19,688,916, unwght n=220) Percent (n)** | **Weighted Percent** | **unweighted** | **p-value** |
| --- | --- | --- | --- | --- | --- | --- | --- |
| *Patient Age Group* | | | | | | | 0.14 |
| 15-24 years | 76,902,789 | 5.21 | 719 | 1,303,095 | 6.62 | 23 |  |
| 25-44 years | 292,120,481 | 19.80 | 2,770 | 5,668,620 | 28.79 | 61 |  |
| 45-64 years | 506,978,063 | 34.36 | 4,432 | 6,957,052 | 35.33 | 71 |  |
| 65-74 years | 317,794,696 | 21.54 | 3,172 | 3,886,335 | 19.74 | 28 |  |
| 75 years and over | 281,841,670 | 19.10 | 2,862 | 1,873,815 | 9.52 | 37 |  |
| Sex-Female | 888,588,434 | 60.22 | 8,078 | 11,011,451 | 55.93 | 122 | 0.49 |
| *Race/Ethnicity* | | | | | | | 0.91 |
| Non-Hispanic White | 1,045,810,087 | 70.87 | 10,561 | 14,721,871 | 74.77 | 159 |  |
| Non-Hispanic Black | 133,530,918 | 9.05 | 975 | 1,603,141 | 8.14 | 21 |  |
| Hispanic | 205,339,297 | 13.92 | 1,673 | 2,230,735 | 11.33 | 21 |  |
| Non-Hispanic Other | 90,957,398 | 6.16 | 746 | 1,133,168 | 5.76 | 19 |  |
| *Source of payment* | | | | | | | N/A |
| All sources blank | 28,178,170 | 1.91 | 328 | 0 | 0.00 | 0 |  |
| Unknown | 40,336,100 | 2.73 | 479 | 421,617 | 2.14 | 2 |  |
| Private insurance | 661,954,793 | 44.86 | 6,147 | 8,862,357 | 45.01 | 86 |  |
| Medicare | 508,458,514 | 34.46 | 5,090 | 6,965,431 | 35.38 | 85 |  |
| Medicaid, CHIP, other state program | 121,603,160 | 8.24 | 1,047 | 1,063,550 | 5.40 | 25 |  |
| Worker's compensation | 37,447,898 | 2.54 | 97 | 73,314 | 0.37 | 1 |  |
| Self-pay | 61,194,737 | 4.15 | 555 | 1,400,468 | 7.11 | 15 |  |
| No charge/charity | 2,422,914 | 0.16 | 28 | 396,318 | 2.01 | 2 |  |
| Other | 14,041,414 | 0.95 | 184 | 505,861 | 2.57 | 4 |  |
| *Is visit related to injury/trauma or adverse effect of medical/surgical treatment* | | | | | | | N/A |
| Blank | 16,256,706 | 1.10 | 173 | 0 | 0.00 | 0 |  |
| Unknown | 62,921,555 | 4.26 | 711 | 41,160 | 0.21 | 9 |  |
| Yes, injury/trauma | 114,425,890 | 7.75 | 800 | 933,734 | 4.74 | 13 |  |
| Yes, overdose/poisoning | 2,918,715 | 0.20 | 11 | 0 | 0.00 | 0 |  |
| Yes, adverse effect of medical/surgical treatment | 17,016,088 | 1.15 | 137 | 219,350 | 1.11 | 4 |  |
| No | 1,254,197,128 | 84.99 | 12,054 | 17,951,024 | 91.17 | 191 |  |
| Questionable injury status | 7,901,617 | 0.54 | 69 | 173,158 | 0.88 | 3 |  |
| *Major reason for visit* | | | | | | | 0.48 |
| Blank | 30,100,566 | 2.04 | 306 | 899,387 | 4.57 | 17 |  |
| New problem (<3 mos. Onset) | 380,438,391 | 25.78 | 3,861 | 3,416,223 | 17.35 | 28 |  |
| Chronic problem, routine | 635,282,040 | 43.05 | 5,789 | 7,848,544 | 39.86 | 84 |  |
| Chronic problem, flare-up | 118,453,798 | 8.03 | 1,366 | 1,912,062 | 9.71 | 35 |  |
| Preventive care | 311,359,904 | 21.10 | 2,633 | 5,612,699 | 28.51 | 56 |  |
| Primary care physician-Yes | 516,980,539 | 35.03 | 3,160 | 13,364,279 | 67.88 | 121 | N/A |
| Established patient-Yes | 1,235,176,226 | 83.70 | 11,223 | 15,991,603 | 81.22 | 193 | 0.66 |
| Does patient have cancer-Yes | 146,532,373 | 9.93 | 1,393 | 1,441,069 | 7.32 | 10 | 0.56 |
| Physician Assistant involved in visit-Yes | 50,182,044 | 3.40 | 511 | 1,358,980 | 6.90 | 4 | 0.17 |
| Nurse Practitioner involved in visit-Yes | 34,765,761 | 2.36 | 208 | 0 | 0.00 | 0 | N/A |
| Other clinician involved in visit-Yes | 425,701,053 | 28.85 | 4260 | 2,471,836 | 12.55 | 29 | 0.01* |
| *Physician specialty* | | | | | | | <0.0001* |
| Primary care | 704,467,590 | 47.74 | 4,383 | 15,073,763 | 77 | 127 |  |
| Surgical care | 291,864,503 | 19.78 | 5,081 | 95,366 | 0 | 3 |  |
| Medical care | 479,305,606 | 32.48 | 4,491 | 4,519,787 | 23 | 90 |  |
| Physician profession-MD | 1,382,956,108 | 93.72 | 1,285 | 16,208,649 | 82 | 147 | <0.05* |
| Solo clinic visit-Yes | 598,176,868 | 40.54 | 5,423 | 11,997,071 | 61 | 136 | <0.05* |
| *Type of clinic ownership* | | | | | | | <0.001* |
| Blank | 37,123,213 | 2.52 | 425 | 2,560,107 | 13 | 12 |  |
| Unknown | 17,430,049 | 1.18 | 108 | 137,632 | 1 | 1 |  |
| Physician/physician group | 1,168,928,292 | 79.22 | 11,259 | 12,397,326 | 63 | 136 |  |
| Medical/academic health center, community health center, other hospital | 89,111,804 | 6 | 731 | 2,242,878 | 11 | 27 |  |
| Insurance company, health plan HMO, other health corporation | 163,044,342 | 11 | 1,432 | 2,350,973 | 12 | 44 |  |
| Rural clinic-Yes | 132,021,007 | 9 | 1,363 | 422,088 | 2 | 5 | 0.005* |
| Alcohol abuse counseling-Yes | 10,145,812 | 1 | 61 | 2,125,678 | 11 | 49 |  |
| Asthma education-Yes | 4,581,729 | 0 | 24 | 623,476 | 3 | 11 |  |
| Ashtma action plan given-Yes | 1,427,686 | 0 | 14 | 310,316 | 2 | 5 |  |
| Diabetes education-Yes | 31,338,471 | 2 | 184 | 2,298,402 | 12 | 26 |  |
| Diet/nutrition-Yes | 157,788,770 | 11 | 1,038 | 12,362,607 | 63 | 134 |  |
| Exercise-Yes | 118,481,708 | 8 | 823 | 11,115,734 | 56 | 128 |  |
| Family planning/contraception-Yes | 15,051,481 | 1 | 128 | 1,247,412 | 6 | 12 |  |
| Genetic counseling-Yes | 8,436,427 | 1 | 33 | 99,845 | 1 | 2 |  |
| Growth/development-Yes | 5,163,952 | 0 | 36 | 120,249 | 1 | 3 |  |
| Injury prevention-Yes | 27,665,309 | 2 | 197 | 3,973,708 | 20 | 70 |  |
| STI prevention-Yes | 6,101,011 | 0 | 34 | 521,294 | 3 | 4 |  |
| Stress management-Yes |  | 0 |  |  | 0 |  |  |
| Substance abuse counseling-Yes | 10,574,762 | 1 | 120 | 2,761,304 | 14 | 57 |  |
| Tobaco use/exposure-Yes | 45,264,367 | 3 | 370 | 3,537,765 | 18 | 59 |  |
| Weight reduction-Yes | 43,224,686 | 3 | 318 | 7,165,194 | 36 | 67 |  |
| Number of medications discussed, Mean (SE) | 3.66 (0.20) | | | 3.74 (0.56) | | | 0.90 |
| Total number of chronic conditions, Mean (SE) | 1.50 (0.09) | | | 2.13 (0.28) | | | 0.03* |
| Number of areas counseled, Mean (SE) | 0.33 (0.03) | | | 2.45 (0.52) | | | <0.0001* |
| Time spent with physician in minutes, Mean (SE) | 23.21 (0.55) | | | 27.84 (3.08) | | | 0.13 |
| Total number of services ordered or provided including vital signs, Mean (SE) | 4.97 (0.16) | | | 9.67 (1.04) | | | <0.0001* |

￼

**Table 2: Descriptive Statistics of Health Education Topics Discussed During Clinic Visits, 2018-2019 (weighted n=1,495,326,615, unweighted=14,175)**

| **Health Education Topic** | **Weighted Frequency (unweighted) All Visits** | **Percent of All Visits (n=14,175)** | **Percent of Primary Care Visits (n=4,520)** | **Visits in which Stress Management was Discussed (n=220)** |
| --- | --- | --- | --- | --- |
| Asthma action plan given | 1,738,002 (19) | 0.12 | 0.24 | 1.58 |
| Asthma education | 5,205,205 (35) | 0.35 | 0.47 | 3.17 |
| Growth/development | 5,284,201 (39) | 0.35 | 0.68 | 0.61 |
| STI prevention | 6,622,305 (38) | 0.44 | 0.91 | 2.65 |
| Genetic counseling | 8,536,271 (35) | 0.57 | 0.14 | 0.51 |
| Alcohol abuse counseling | 12,271,490 (110) | 0.82 | 0.87 | 10.8 |
| Substance abuse counseling | 13,336,066 (177) | 0.89 | 1.21 | 14.02 |
| Family planning/contraception | 16,298,893 (140) | 1.09 | 2.04 | 6.34 |
| Stress management | 19,688,916 (220) | 1.32 | 2.09 | - |
| Injury prevention | 31,639,017 (267) | 2.12 | 2.25 | 20.18 |
| Diabetes education | 33,636,873 (210) | 2.25 | 3.69 | 11.67 |
| Tobacco use/exposure | 48,802,132 (429) | 3.26 | 4.57 | 17.97 |
| Weight reduction | 50,389,880 (385) | 3.37 | 4.61 | 36.39 |
| Exercise | 129,597,442 (951) | 8.67 | 13.95 | 56.46 |
| Diet/nutrition | 170,151,377 (1,172) | 11.38 | 17.26 | 62.79 |
| Visits in which at least one area was discussed | 279,518,938 (2,076) | 18.69 | 25.78 | - |
| Number areas discussed-mean (SE) | 0.36 (0.03) | | 0.53 (0.05) | 2.45 (0.24) |

**Table 3: Weighted logistic regression predicting whether stress was discussed during clinic visit (weighted n=1,495,326,615, unweighted=14,175)**

| **Independent Variable** | **Odds Ratio (OR)** | **95% CI** |
| --- | --- | --- |
| Other clinician involved in visit-Yes | 0.30 | 0.12-0.77* |
| *Physician Specialty* | | |
| Primary care | Reference | |
| Medical care specialist | 0.87 | 0.36-2.13 |
| Surgical care specialist | 0.06 | 0.01-0.29* |
| Doctor of Medicine-Yes | 1.38 | 0.32-5.94 |
| Solo clinic | 3.29 | 1.39-7.81* |
| *Type of health system* | | |
| Medical/academic health center, community health center, other hospital | Reference | |
| Blank | 4.03 | 1.29-12.61* |
| Insurance company, health plan, HMO, other health corporation | 0.37 | 0.11-1.27 |
| Physician or physician group | 0.23 | 0.09-0.59 |
| Unknown | 0.09 | 0.01-1.52 |
| Rural area-Yes | 0.22 | 0.09-0.53* |
| Number areas counseled during visit | 2.10 | 1.60-2.75 |
| Depression screening conducted during visit-Yes | 1.70 | 0.59-4.89 |
| Number of services ordered or provided during visit | 1.10 | 1.02-1.18 |
| Total number of chronic conditions | 1.11 | 0.92-1.33 |
| Patient age | 0.99 | 0.96-1.01 |

*Statistically significant at p<0.05
